# *CardioBrain*: A Novel User-Friendly Software for Assessing Dynamic Cerebral Autoregulation and Cardiovascular Interactions

**DOI:** 10.1101/2024.10.21.24315862

**Authors:** Tiago Pecanha, Rhenan Bartels, Gabriel Dias Rodrigues

**Affiliations:** Department of Sport and Exercise Sciences, Manchester Metropolitan University Institute of Sport, Manchester Metropolitan University, Manchester, UK; Independent researcher. Software engineer at Global Minimum LTDA. Rio de Janeiro, Brazil; Department of Physiology and Pharmacology, Fluminense Federal University, Niterói, RJ, Brazil; Department of Clinical Sciences and Community Health, University of Milan, Milan, Italy

**Keywords:** brain vasculature, power spectral density, middle cerebral artery, cerebrovascular control, transcranial doppler

## Abstract

Cerebral autoregulation (CA) is a critical mechanism that maintains cerebral blood flow (CBF) relatively stable despite fluctuations in arterial blood pressure (ABP), ensuring protection against ischemia and hyperperfusion. Alterations in CA are linked to adverse outcomes in various conditions, highlighting the need for precise and accessible methods to analyse CA. *CardioBrain* is a freely available user-friendly software developed to assess dynamic CA by performing Transfer Function Analysis (TFA) on continuously recorded ABP and CBF data. The software utilizes the Welch method for spectral density and cross-spectral analysis. Following TFA, the software calculates different dCA metrics as gain, phase, and coherence across various frequency bands (VLF, LF, HF). *CardioBrain* allows users to easily manipulate signal processing parameters, visualize data, and export results, making it suitable for both research and clinical settings without requiring advanced programming skills. Additionally, the software includes tools to address signal misalignments, such as a temporal shift feature, and ensures adherence to recommended standards for TFA analysis. Future development will focus on incorporating user feedback, validating the software against simulated and real-world data, and expanding its functionality for a broader range of regulatory analyses.

## Introduction

Cerebral autoregulation (CA) represents the ability of the brain’s arteries and arterioles to maintain cerebral perfusion despite fluctuations in arterial blood pressure (ABP) (1, 2). CA encompasses multiple regulation mechanisms (e.g., myogenic control as well as metabolic and neurogenic mechanisms) (1, 3), which work in concert to ensure relatively stable cerebral blood flow (CBF) across a wide range of ABP, protecting against both ischemia and hyperperfusion. This mechanism may protect the brain by safeguarding neuronal function, maintaining intracranial pressure, and reducing the risk of brain damage due to arterial blood pressure (ABP) fluctuations. (2, 4, 5).

Alterations of CA have been described and linked with poor outcomes in a range of conditions, including in hypertension (6), heart failure (7), atrial fibrillation (8), stroke (9), traumatic brain injury (10, 11), and in patients under intensive care (5, 12), which further highlights the clinical significance of this mechanism as well as the necessity of developing applications to quantify CA using continuous physiological data.

The approaches used to investigate cerebral autoregulation (CA) can be divided by their methodology and timescale and are typically classified as static or dynamic (1, 2). Static CA involves assessing the relationship between ABP and CBF under steady-state conditions over longer time periods (usually >10 minutes). This typically involves experiments where ABP is manipulated—either increased or decreased—to determine the range over which CBF remains stable (2). Alternatively, dynamic CA (dCA) can be studied by promoting rapid changes in ABP and measuring how quickly and effectively the cerebral vasculature adapts to dampen potential fluctuations in CBF (1, 2). Different protocols have been developed to induce rapid changes in ABP to assess dCA, including the squat-to-stand manoeuvre (13, 14), oscillatory lower body negative pressure (15), respiratory manoeuvres (e.g., Valsalva or paced breathing) (16, 17), the tight cuff inflation-deflation manoeuvre (3), and others. The relationship between ABP and CBF during these tests can be analysed using many methodologies, including Transfer Function Analysis (TFA), which allows for the assessment of gain, phase, and coherence between ABP and CBF fluctuations, providing detailed insights into the dynamic characteristics of CA.

Over the last 13 years, the Cerebrovascular Research Network (CARNet) has gathered interest and organized a joint effort to set the standards for assessing dCA using TFA (18, 19). The publication of the 2016 and 2022 White Papers has improved consistency in the collection, processing, and analysis of CA-related data using TFA. However, the relative complexity of the analysis process, coupled with the lack of accessible, user-friendly software solutions, remains a significant barrier to wider adoption. A user-friendly software solution could help overcome this challenge by automating data processing and analysis, ensuring adherence to standardized guidelines, and making TFA more accessible to clinicians and researchers without advanced coding expertise.

This paper presents and marks the release of *CardioBrain*, a user-friendly software designed for the analysis of dCA using continuously obtained ABP and CBF data through the TFA. Here, we have highlighted the main features of the software, introducing a tool that simplifies data processing and makes cerebral autoregulation analysis more accessible to a broader range of users.

## Implementation

### Data Processing in CardioBrain: Spectral and Transfer Function Analysis

The CardioBrain software performs spectral density analysis of ABP (input) and CBFV (output) using the Welch method (for more information: (20)). This method involves dividing the data into overlapping segments, applying a window function to each segment to reduce spectral leakage, and averaging the periodograms obtained from the Fast Fourier Transform (FFT) of each segment. The software calculates the power spectral density (PSD) for ABP and CBFV separately, generating their respective auto-spectra. Following this, the software computes the cross-spectra by multiplying the complex conjugate of the FFT of ABP with the FFT of CBFV. This cross-spectral analysis highlights the relationship between the two signals in terms of phase and magnitude across different frequency bands. Finally, the software conducts transfer function analysis (TFA) by dividing the cross-spectra by the auto-spectra of ABP. This process allows the estimation of key metrics such as gain, phase, and coherence, which are reported as the primary outcomes for cerebral autoregulation (CA) analysis.

A simplified formula for Transfer Function Analysis (TFA) can be expressed as:

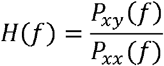

Where:

- H(*f*) is the transfer function at frequency f,
- P_xy_(*f*) is the cross-spectrum between the input signal (ABP) and the output signal (CBFV),
- P_xx_(*f*) is the auto-spectrum (power spectral density) of the input signal (ABP).

### Software development and compatibility

CardioBrain was developed in Python and is a standalone multi-platform executable (.exe) developed for compatibility with Windows, macOS, and Linux systems.

#### Data Input Format

The software processes data in CSV file format, specifically designed for time series analysis. Each file must follow a structured format, where the first column contains beat-by-beat RR intervals (RRi) in seconds, the second column represents time-averaged mean cerebral blood flow velocity (CBFV) in cm/s, and the third column contains mean arterial blood pressure values in mmHg (ABP) (Figure 1). It’s important to note that the order of these columns must not be changed, as the software expects this exact sequence for proper data processing. Additionally, the software is designed to ignore the headers in the first row, so this can be used to describe the appropriate column headers “RRi,” “CBFV,” and “ABP as demonstrated in Figure 1.” However, if a second row contains text data, the software will generate an error message: “Error. Could not open file,” as it requires numeric data starting from the second row onward.

**Figure 1.**
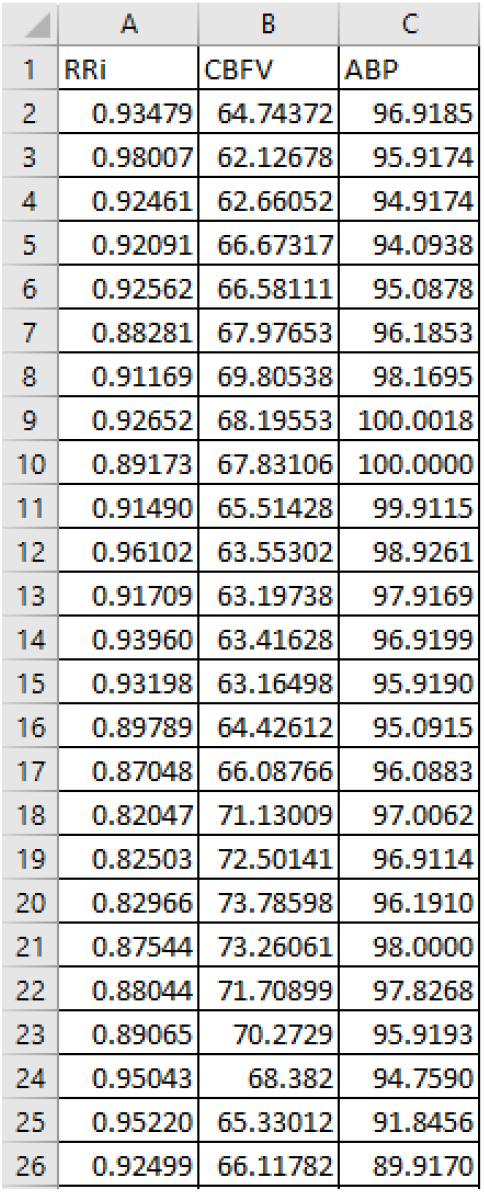
The CSV file must include the required inputs in the following order: RRi (RR interval in seconds), CBFV (cerebral blood flow velocity in cm/s), and ABP (mean arterial blood pressure in mmHg). The columns must remain in this specific sequence, as the software depends on this arrangement for accurate data processing.

### Software interface

Figure 2 presents the software interface, including the Control Panel (Fig 2A), the Graphical Interface (Fig 2B), and the Results Table (Fig 2C). The Control Panel allows users to configure various signal processing parameters in line with the recommendations from the CARNet (18, 19). The Graphical Interface displays synchronized time-series plots of the CBFV and ABP signals. The Results Table provides relevant outcomes from the TFA, such as gain, coherence, phase, and power across different frequency bands for both ABP and CBFV signals.

**Figure 2.**
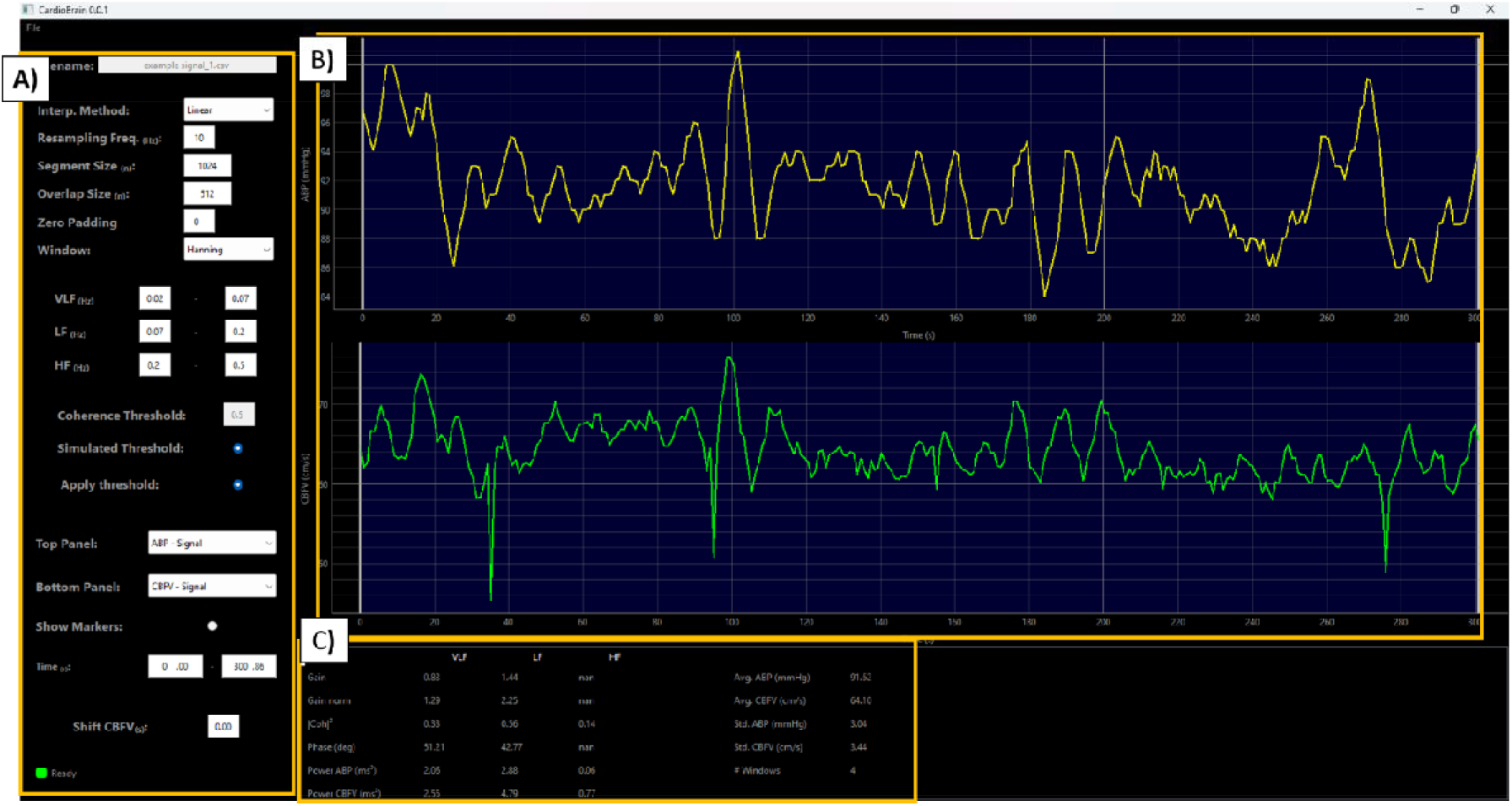
Overview of the *CardioBrain* software interface, showing the Control Panel (Panel A), the Graphical Interface (Panel B), and the Results Table (Panel C).

### Graphical interface

Once a file is opened in the software, the CBFV and ABP time series are plotted synchronously in the graphical interface. The ABP (Arterial Blood Pressure) signal is displayed in the top graph, while the CBFV (Cerebral Blood Flow Velocity) signal is shown in the bottom graph.

The graphical interface (Figure 3) brings important features related to signal display and selection. The interface includes vertical lines at each extremity of the time series plots, which are used to select the specific portion of the signal to be analysed. These lines can be moved along the timeline to define the ‘analysis segment’. The analysis segment is highlighted in blue as demonstrated in Figure 3. The CBFV and ABP signals are synchronized, meaning that when a user adjusts the selection on one signal, the corresponding selection on the other signal will move simultaneously. This ensures that both signals are analysed over the same time segment, maintaining their temporal alignment for accurate TFA. By right-clicking on the plot, users can access additional options to customize the display, including manually adjusting the range for each axis and plotting transformed signals, such as logarithmic values or power spectral density (PSD), for more detailed analysis (Figure 3).

**Figure 3.**
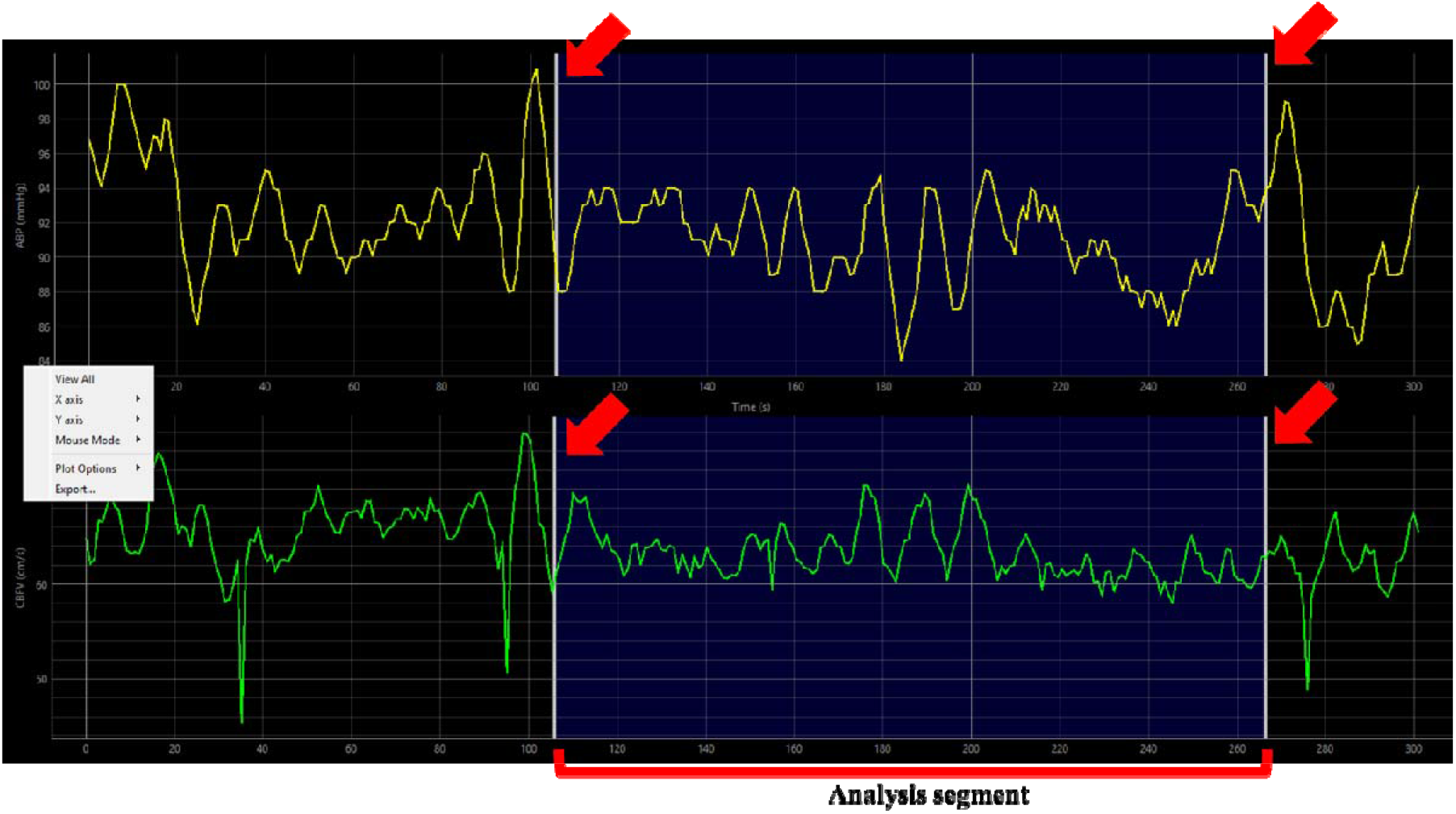
Graphical interface. In this example, the segment that is being analysed is highlighted in blueand marked by a red horizontal bracket, and goes from ∼110s to ∼270s. The analysis window can be selected by dragging the white vertical lines (indicated by the red arrows) sideways. Right-clicking the mouse opens a menu that allows users to modify the Y and X axes’ range (scale) and access additional plot options. The option “View All” is a useful feature that resets each signal to its automatic scale.

### Control panel

The software employs the Welch method to calculate the averaged auto- and cross-spectra of each signal, which are then used for TFA. The Control Panel (Fig 2A) allows users to adjust key signal processing parameters that influence the Welch analysis, including the interpolation method, resampling frequency, segment size (or window length in data points), overlap size (to determine the superposition between window segments), zero padding, and the type of anti-leakage window (e.g., Hanning, Hamming). The adjustable parameters offer flexibility in how the data is analysed. For example, the number of data points in the segment size, combined with the resampling frequency, can be used to determine an optimal window length in seconds (e.g., a resampling frequency of 10 Hz and a segment size of 1024 points results in a window length of 102.4 seconds). The interface also allows for the customization of the frequency range across standard frequency bands—VLF (Very Low Frequency, typically 0.02-0.07 Hz), LF (Low Frequency, typically 0.07-0.20 Hz), and HF (High Frequency, typically 0.20-0.50 Hz). All the above parameters and their underlying options were selected in line with the recommended settings for TFA as proposed by the 2016 and 2022 White Papers from the CARNet (18, 19).

The software also has some fixed processing parameters that are built into the main algorithm and are not offered to the user as adjustable options, simplifying the analysis process for the user. For instance, the removal of mean values from both CBFV and ABP signals is performed automatically before FFT analysis. Additionally, both auto- and cross-spectra are smoothed using a triangular window with fixed coefficients of 0.25, 0.5 and 0.25 prior to the transfer function calculation.

#### Graphical display tool

In the Control Panel, the software includes a graphical display feature (Figure 4) that allows users to display various plots derived from the PSD and TFA analyses in the top and bottom panels of the graphical interface (Fig 4A). These plots include options such as PSD (Fig 4B), gain, phase, and coherence. The software also provides an option to plot both ABP and CBFV in the same panel (ABP x CBFV, Fig 4C), which can be useful for evaluating the alignment and temporal relationship between the signals. The resulting graphs can be exported as image files by right-clicking and selecting the export option (Fig 4D), facilitating their inclusion in papers and presentations.

**Figure 4.**
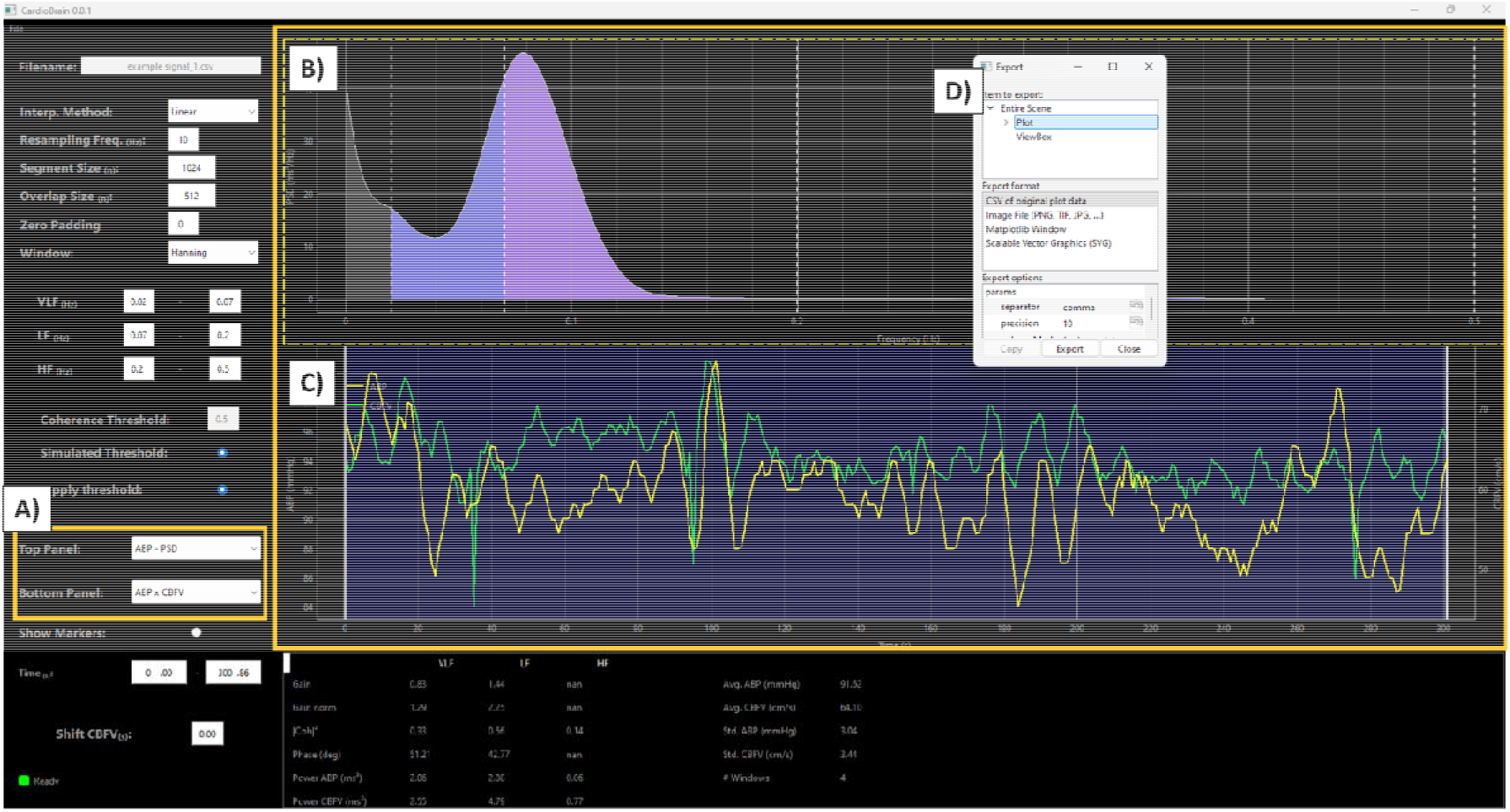
Users can select different parameters of the TFA analysis to be displayed in the graphical interface (using the graphical display tool in Panel A), including the original ABP and CBFV signals, their respective power spectral densities (PSD), and gain, phase, and coherence estimates from the transfer function analysis (TFA) across all frequency bands of the cross-spectrum. Panel B displays the PSD of the ABP signal, while Panel C shows both the ABP and CBFV time series in the same plot, highlighting their temporal alignment. Either or both panels can be exported as figure files by right-clicking and choosing the desired format (Panel D).

**Figure 5.**
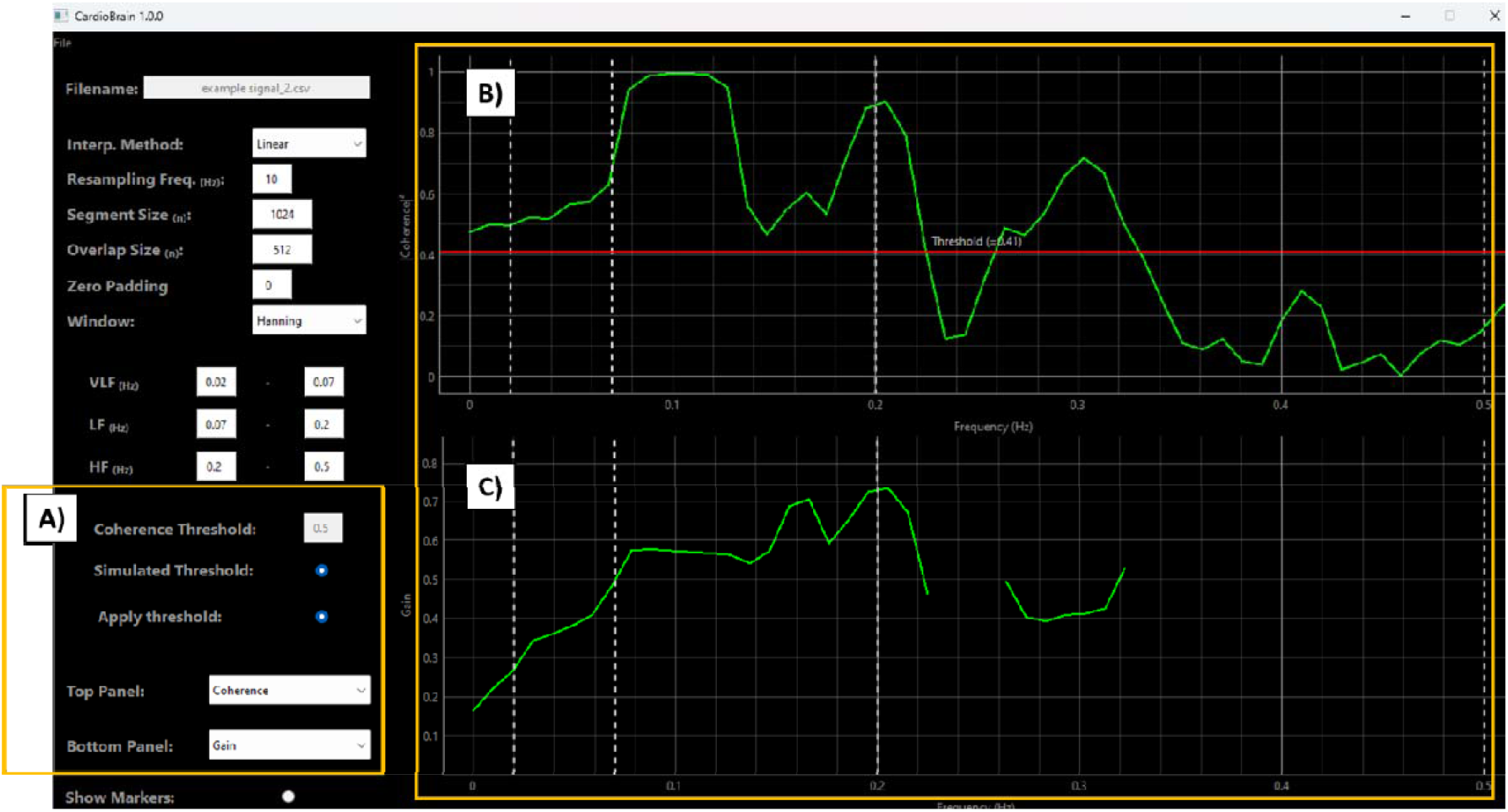
Control Panel and Coherence Function Analysis. The control panel (panel A) provides two options for the user to establish the critical squared coherence for TFA. Under ‘Coherence Threshold’, the user can set a fixed predefined threshold (e.g., 0.5). Alternatively, users can select the Simulated Threshold option, which prompts the software to use the cut-off values for coherence (i.e., critical values of coherence at □=0.05) provided by the CARNet White Paper 2022 (19). The software also allows users to verify the applied threshold by plotting the squared coherence across all frequency bands in the top panel of the graphical display, and either the gain or phase in the bottom panel. In the example shown, the top panel (Panel B) illustrates the values of squared coherence across all frequency bands. In this example, the software simulated a critical value for the squared coherence of 0.4, which is marked by the red horizontal line in the top panel. The bottom panel (Panel C) shows the estimated gain, but only for the portions of the spectrum where the squared coherence surpassed the designated threshold. This ensures that only portions of the spectrum meeting or exceeding the threshold are used to estimate gain, phase, or other TFA-related measures. The user can also deselect the ‘Apply Threshold’ option, which will deactivate the coherence-based analysis, allowing the software to estimate TFA-related outcomes using the entire spectrum.

#### Coherence function

The coherence function tool is an essential feature that ensures the validity of the gain and phase estimates from the TFA. The software provides two options for coherence function analysis. Under the Coherence Threshold option, the user can set a fixed threshold (or cut-off) value for the squared-coherence function to ‘accept’ the validity of estimates of gain and phase at each frequency (e.g., 0.5). Alternatively, the user can select the Simulated Threshold option, which utilizes the cut-off values for coherence (i.e., critical values of coherence at □ =0.05) provided by the CARNet White Paper 2022 (19). This approach is based on Monte Carlo simulations and utilizes the number of windows from the Welch method. Typically, a larger number of windows lowers the critical value for squared coherence.

After applying the selected threshold—whether a fixed coherence threshold or a simulated one— the software automatically accepts only those gain and phase estimates that exceed the designated threshold. This step ensures that only reliable estimates are used for analysis, reducing the influence of noise or unreliable frequency bands. The user can also deselect the ‘Apply Threshold’ option, which disables the coherence-based analysis and enables the software to estimate TFA-related outcomes across the full spectrum.

The software provides an option for the user to verify the applied threshold and identify which portions of the cross-spectrum were used to estimate gain and phase based on achieving the designated critical threshold. This can be done by plotting the coherence in the top panel and either the phase or gain in the bottom panel, using the options available in the Control Panel. This allows the user to visually inspect the coherence and corresponding gain or phase values across the frequency range, ensuring that only regions of the cross-spectrum exceeding the critical threshold are used for further analysis.

#### Temporal shift tool

The *CardioBrain* software includes a signal shift tool (Figure 6) designed to address temporal misalignments between ABP and CBFV signals, which can arise due to integration delays in recording devices. In some physiological monitoring systems, devices that measure ABP or CBFV may introduce small time delays, resulting in an offset between the two signals. This misalignment can affect the accuracy of the TFA and impact the estimations of gain, phase, and coherence, which are critical for analysing dCA.

**Figure 6.**
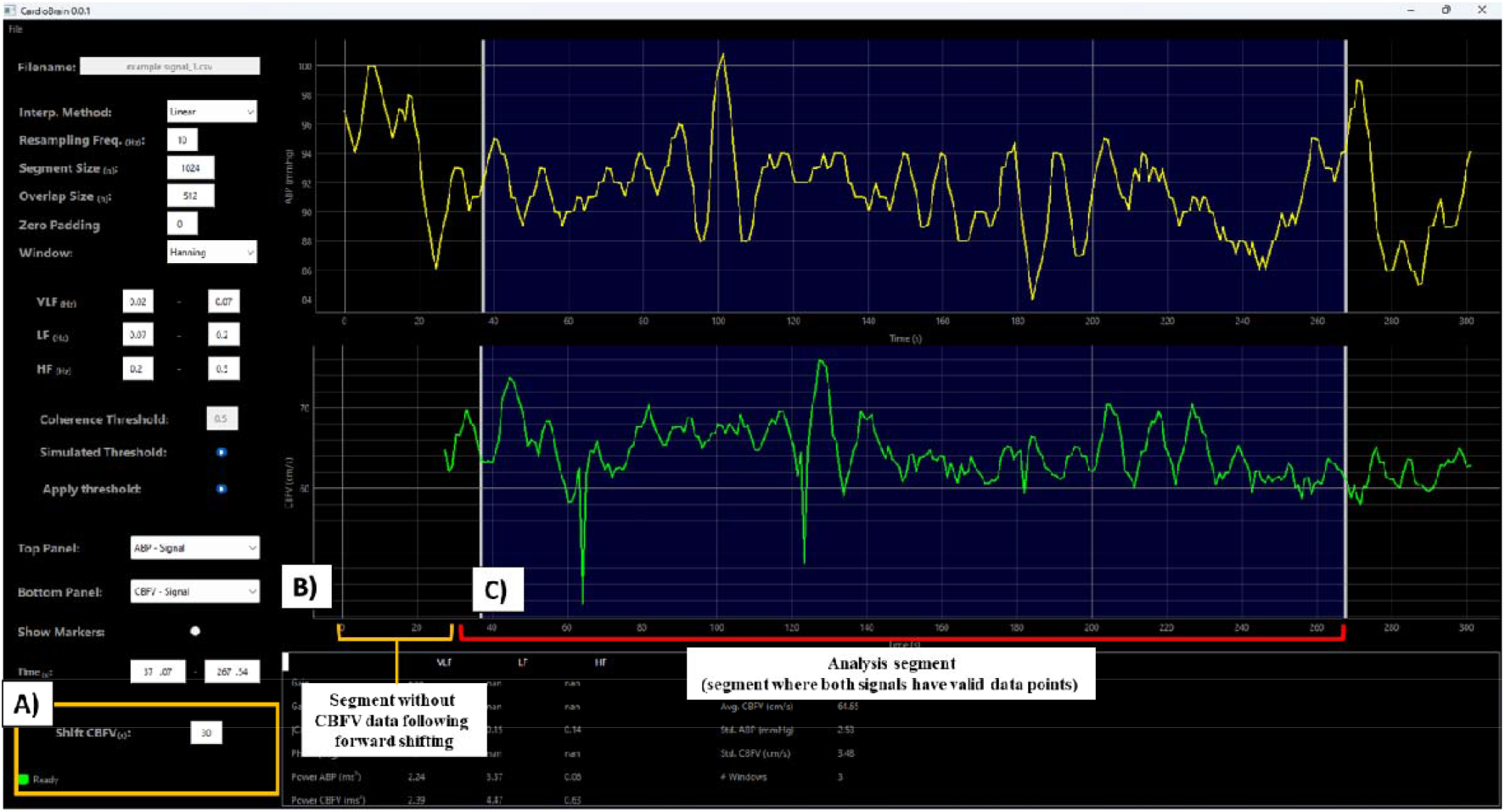
Temporal Shift Tool. This tool allows users to manually adjust the CBFV signal forward or backward in time by a specified number of seconds (use positive values to shift forward and negative values to shift backward) (Panel A). Following temporal shifting, there will be a segment of the CBFV signal with missing data, either at the beginning for forward shifts or at the end for backward shifts (Panel B). Users must then drag the vertical selection lines to a region where both signals, ABP and CBFV, contain valid data for accurate analysis (as demonstrated in Panel C).

To correct this, the signal shift tool allows users to manually adjust the CBFV signal forward or backward in time by a specified number of seconds (use positive values to shift forward and negative values to shift backward) (Fig 6A). Importantly, this shift will create a section of the CBFV signal without data—either at the beginning if the signal is shifted forward or at the end if it is shifted backward (Fig 6B). To ensure proper analysis, users must adjust the vertical selection lines to encompass a section of the time range where both signals (ABP and CBFV) have valid data points (Fig 6C). This ensures that the TFA computations are based on properly aligned segments, preventing errors in gain, phase, and coherence analysis due to missing data.

### Results interface

Figure 7 depicts the results interface of the *CardioBrain* software. The software automatically generates all results as the user selects and updates parameters in the Control Panel and adjusts the analysis segment using the vertical selection lines. Following the recommendations from the CARNet White Papers (18, 19), the software calculates and reports the mean values of gain, phase, and coherence for the TFA across the VLF, LF, and HF frequency bands. The software calculates gain in absolute units (Gain; in cm.s^-1^.mmHg^-1^) or normalised by the mean CBFV (Gain norm; in %.mmHg^-1^). Additionally, the software reports the PSD across all frequency bands for both the ABP and CBFV signals, as well as the mean and standard deviation for each of these signals. Furthermore, the software provides detailed information regarding the number of windows generated during the Welch analysis, which is influenced by the user-specified resampling frequency, segment size, and overlap size.

**Figure 7.**
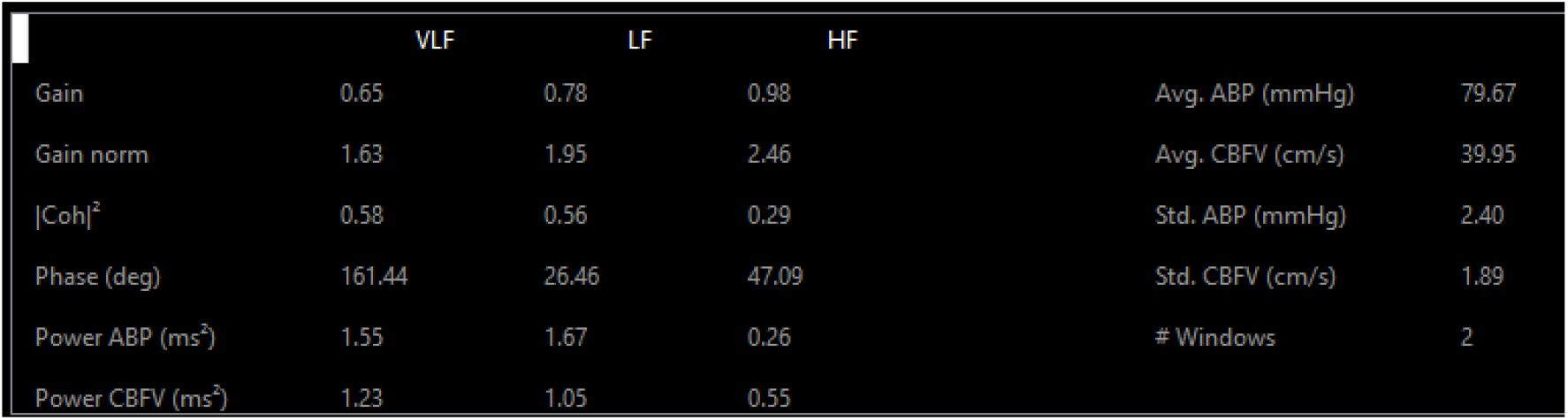
Results table generated by the *CardioBrain* software, presenting key parameters from the TFA and PSD analyses. The table reports values for absolute gain (Gain), normalized gain (Gain norm), coherence (|Coh|^2^), phase, and power for ABP and CBFV across the VLF, LF, and HF frequency bands. Additionally, it provides the mean and standard deviation of ABP and CBFV, along with the number of windows used in the Welch analysis, which in the above example is 2.

After the data analysis is complete, users can save/export the results via the File menu, which generates a CSV file containing all the calculated parameters displayed in the results interface, enabling further analysis or reporting.

## Conclusion and next steps of development

This paper presented the key features of *CardioBrain*, a user-friendly software designed for the analysis of dCA using TFA. *CardioBrain* is not only intuitive and easy to use, but it is also freely available, operating independently of programming environments like Python or MATLAB, making it accessible to a broad range of users, including those without advanced coding skills. Importantly, the software provides detailed metrics, including gain, phase, and coherence across all relevant frequency ranges, making it a valuable tool for the assessment of CA, particularly in datasets obtained from continuous ABP and CBFV recordings. In addition to dCA analysis, the software also reports variability in ABP and CBFV across physiologically relevant frequencies, which could be used to further explore the regulation of both ABP and CBFV. Relevant for both dCA and broader cardiovascular regulatory analyses, the software’s tools and functionalities allow for the calculation of all relevant PSD and TFA parameters during spontaneous resting recordings of ABP and CBFV, as well as during protocols designed to induce forced oscillations at a fixed frequency (e.g., 0.10 Hz repeated squat-stands).

This paper marks the release of *CardioBrain Version 1.0. CardioBrain* is freely available for both academic and clinical use and can be requested using the. The next steps of development include the implementation of additional features, which will be informed by user feedback from the research and clinical communities using the software. Additionally, in a forthcoming publication, we will test the validity of the software against simulated models and real-world data to further ensure its accuracy and robustness for widespread use.

## Data Availability

All data produced in the present study are available upon reasonable request to the authors

